# The COVID-19 pandemic storm in India

**DOI:** 10.1101/2021.05.06.21256523

**Authors:** Igor Nesteruk

## Abstract

The sharp increase in the number of new COVID-19 patients in India in the second half of April 2021 has caused alarm around the world. A detailed analysis of this pandemic storm is still ahead. We present the results of anterior analysis using a generalized SIR-model (susceptible-infected-removed). The final size of this pandemic wave and its duration are predicted. Obtained results show that the COVID-19 pandemic will be a problem for mankind for a very long time.

## Introduction

The daily number of new laboratory-confirmed COVID-19 cases in India exceeded 400,000 in the end of April 2021. This huge figure is frightening, but if we take into account the population of India, the number of new cases per capita is not yet higher than the maximum for some other countries, including Ukraine. It is very important to assess the growing trends in the number of new cases and the ability of the Indian medical system to cope with the huge number of patients and deaths.

Any mathematical modeling of the epidemic dynamics will be of particular value if we make an accurate long-term forecast of its duration and number of diseases using statistics data sets obtained immediately after the outbreak. Many authors are trying to predict the Covid-19 pandemic dynamics in many countries and regions [1-72]. We will not analyze these studies in details and only note that the correct mathematical simulation of the Covid-19 pandemic is very difficult for at least two reasons.

First, data on the number of cases are clearly incomplete immediately after the epidemic outbreak, there are quite long hidden periods [67, 70, 73-77]. In particular, first COVID-19 cases probably have appeared already in August 2019 [67, 70]. The reason is the large number of asymptomatic patients and the lack of skills to detect a new disease. It must be noted that large discrepancy between registered and actual number of cases occurred even for later periods of Covid-19 pandemic [72, 78-80].

The second reason for the limited accuracy of long-term forecasts is the constant changes in the conditions of the pandemic (quarantine measures, social behavior, virulence of the pathogen, etc.). Therefore, a prediction made using statistics for a certain time period is not suitable for other periods of time. To solve this problem a generalized SIR-model and the methods of its parameter identification was proposed in [70, 81, 82]. Since the new pandemic wave in India is not the first one we will use the generalized SIR-model and the method of direct parameter identification (without calculations of the previous epidemic waves) [83]. Corresponding results for the first Covid-19 epidemic waves in mainland China, USA, Germany, the UK, the Republic of Korea, Austria, Italy, Spain, France, the Republic of Moldova, Qatar, Ukraine, the city of Kyiv and for the world are already published in [65, 67, 69-72, 81, 82] and showed rather good accuracy. In this article we will apply the same approach to the case of India.

### Data

We will use the data set regarding the accumulated numbers of confirmed COVID-19 cases in India (*V*_*j*_) from Data Repository by the Center for Systems Science and Engineering (CSSE) at Johns Hopkins University (JHU) [84]. The values *V*_*j*_ and corresponding time moments *t*_*j*_ (measured in days) are shown in Table 1.

**Table 1.**
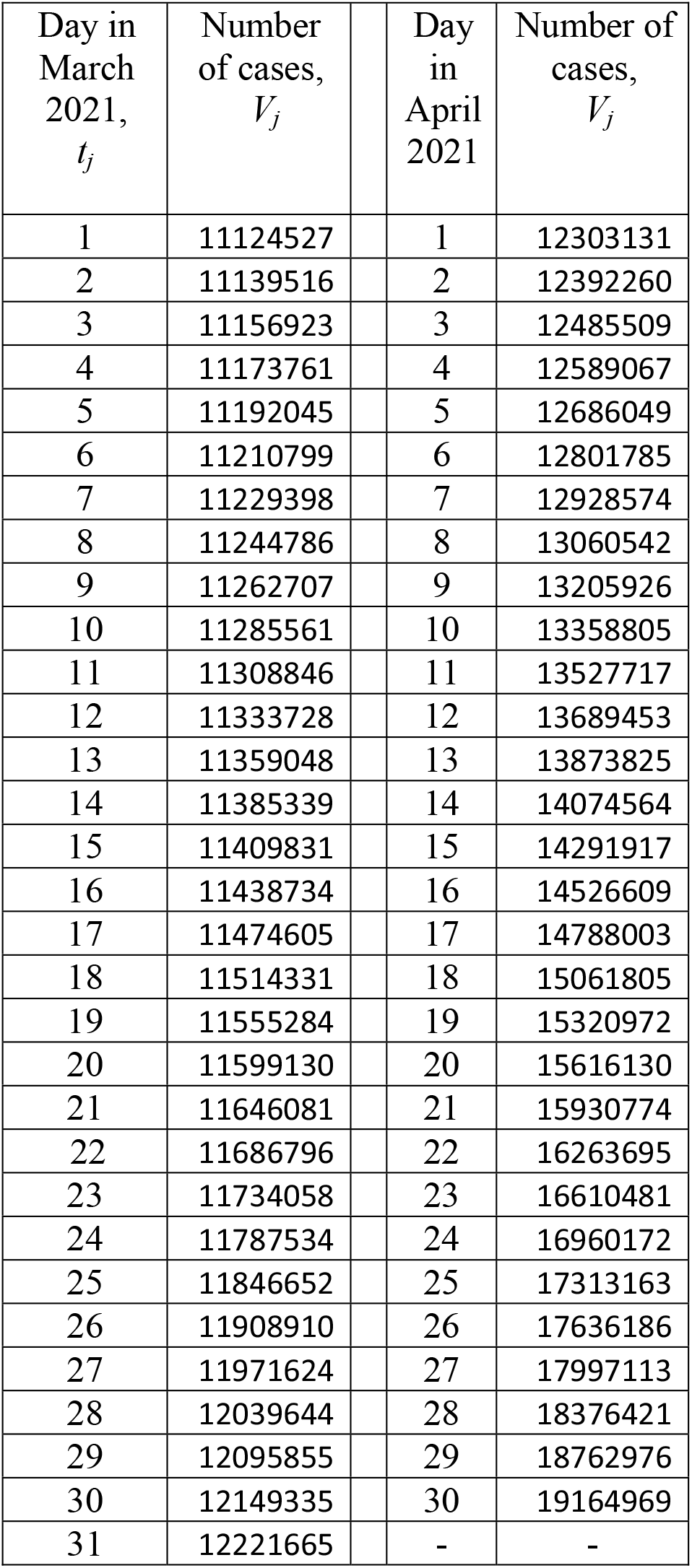
**Cumulative numbers of laboratory confirmed Covid-19 cases in India *V*_*j*_ according to JHU, [84].**

| Day in March 2021, $t_j$ | Number of cases, $V_j$ | | Day in April 2021 | Number of cases, $V_j$ |
| --- | --- | --- | --- | --- |
| 1 | 11124527 |  | 1 | 12303131 |
| 2 | 11139516 |  | 2 | 12392260 |
| 3 | 11156923 |  | 3 | 12485509 |
| 4 | 11173761 |  | 4 | 12589067 |
| 5 | 11192045 |  | 5 | 12686049 |
| 6 | 11210799 |  | 6 | 12801785 |
| 7 | 11229398 |  | 7 | 12928574 |
| 8 | 11244786 |  | 8 | 13060542 |
| 9 | 11262707 |  | 9 | 13205926 |
| 10 | 11285561 |  | 10 | 13358805 |
| 11 | 11308846 |  | 11 | 13527717 |
| 12 | 11333728 |  | 12 | 13689453 |
| 13 | 11359048 |  | 13 | 13873825 |
| 14 | 11385339 |  | 14 | 14074564 |
| 15 | 11409831 |  | 15 | 14291917 |
| 16 | 11438734 |  | 16 | 14526609 |
| 17 | 11474605 |  | 17 | 14788003 |
| 18 | 11514331 |  | 18 | 15061805 |
| 19 | 11555284 |  | 19 | 15320972 |
| 20 | 11599130 |  | 20 | 15616130 |
| 21 | 11646081 |  | 21 | 15930774 |
| 22 | 11686796 |  | 22 | 16263695 |
| 23 | 11734058 |  | 23 | 16610481 |
| 24 | 11787534 |  | 24 | 16960172 |
| 25 | 11846652 |  | 25 | 17313163 |
| 26 | 11908910 |  | 26 | 17636186 |
| 27 | 11971624 |  | 27 | 17997113 |
| 28 | 12039644 |  | 28 | 18376421 |
| 29 | 12095855 |  | 29 | 18762976 |
| 30 | 12149335 |  | 30 | 19164969 |
| 31 | 12221665 |  | - | - |

### Generalized SIR model and parameter identification procedure

The classical SIR model for an infectious disease [85-87] was generalized in [70, 72, 81] to simulate different epidemic waves. We suppose that the SIR model parameters are constant for every epidemic wave, i.e. for the time periods: 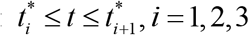. Than for every wave we can use the equations, similar to [85-87] :

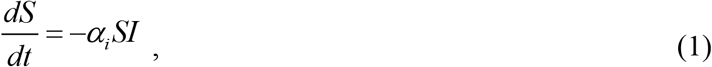

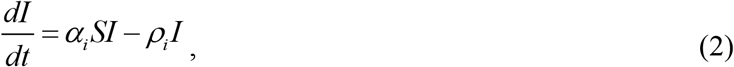

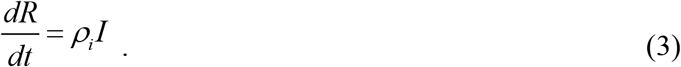

Here *S* is the number of susceptible persons (who are sensitive to the pathogen and **not protected**); *I* is the number of infectious persons (who are sick and **spread the infection)**; and *R* is the number of removed persons (who **no longer spread the infection)**. It must be noted that *I(t)* is not the number of active cases. People can be ill (among active cases), but isolated. In means, that they don’t spread the infection anymore. There are many people spreading the infection but not tested and registered as active cases. The use of number of active cases as *I(t)* in some papers a principal mistake which may lead to incorrect results. Parameters α_*i*_ and ρ_*i*_ are supposed to be constant for every epidemic wave.

To determine the initial conditions for the set of equations (1)–(3), let us suppose that at the beginning of every epidemic wave 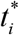 :

In [70, 72, 81] the set of differential equations (1)-(3) was solved with the use of initial conditions

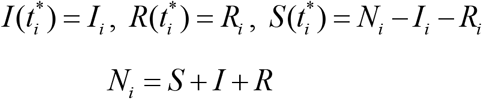

and by introducing the function *V* (*t*) = *I* (*t*) + *R*(*t*), corresponding to the number of victims or the cumulative confirmed number of cases. For many epidemics (including the COVID-19 pandemic) we cannot observe dependencies *S*(*t*), *I* (*t*) and *R*(*t*) but observations of the accumulated number of cases *V*_*j*_ corresponding to the moments of time *t*_*j*_ provide information for direct assessments of the dependence *V* (*t*). The corresponding analytical formulas for this exact solution; the saturation levels *S*_*i*#x221E;_ ; *V*_*i*∞_ = *N*_*i*_ − *S*_*i*∞_ (corresponding the infinite time moment) and the final day of the *i-th* epidemic wave (corresponding the moment of time when the number of persons spreading the infection will be less than 1) can be found in [70, 72, 81].

The exact solution depends on five parameters - *N*_*i*_, *I*_*i*_, *R*_*i*_,*ν*_*i*_,α_*i*_. The values *V*_*j*_, corresponding to the moments of time *t*_*j*_ can be used to find the optimal values of this parameters corresponding to the maximum value of the correlation coefficient *r*_*i*_ [88]. The details of this approach can be found in [89, 90]. It was successfully used in [65, 67, 69-72, 81-83, 89, 91, 92] to simulate the COVID-19 pandemic dynamics and other phenomena. The exact solution [70, 72, 81] allows avoiding numerical solutions of differential equations (1)-(3) and significantly reduces the time spent on calculations. The new algorithm proposed in [90] allows estimating the optimal values of SIR parameters for the *i-th* epidemic wave directly (without simulations of the previous waves) with the use of only two independent parameters *N*_*i*_ and *ν*_*i*_.

## Results

The optimal values of parameters and other characteristics of the severe COVID-19 pandemic wave in India are calculated and listed in Table 2. We have used the number *i=2* and the time period: *T*_*c2*_ - April 10-23, 2021 for SIR simulations of this wave. Corresponding values of *V*_*j*_ and *t*_*j*_ are listed in Table 1. The value of the correlation coefficient *r*_*i*_ = 0.999959712033103 is very high (see Table 2), nevertheless we are not satisfied with the convergence procedure by isolation of the maximum of this parameter. Probably new simulation with the use of fresher datasets could fix this problem. The estimate of the average duration of the infection spread in India *τ*_*i*_ = 1/ ρ_*i*_ ≈ 35 days is significantly higher than in Ukraine in the end of March 2021, [72].

**Table 2.** **The COVID-19 pandemic storm in India. Optimal values of SIR parameters and other characteristics.**

| Characteristics | $i=2$ |
| --- | --- |
| Time period taken for calculations $T_{ci}$ | April 10-23, 2021 |
| $I_i$ | 1,388,033.10466357 |
| $R_i$ | 11,836,652.8953364 |
| $N_i$ | 43,000,000 |
| $\nu_i$ | 7,965,355.17869963 |
| $\alpha_i$ | 3.58639081028928e-09 |
| $\rho_i$ | 0.0285668766135785 |
| $1/\rho_i$ | 35.0055770368917 |
| $r_i$ | 0.999959712033103 |
| $S_{i\infty}$ | 645,561 |
| $V_{i\infty}$ | 42,354,439 |
| Final day of the epidemic wave | June 23, 2033 |

Unfortunately the estimations of the pandemic duration in India are very pessimistic (4,434 days or 12.1 years after April 30, 2021). If we suppose that the end of the epidemic corresponds the moment when the number of persons spreading the infection is less than 20, the calculations yield the middle of January 2023. Probably a strict quarantine and vaccination could change this sad trend, but it looks that some new cases of COVID-19 will appear if not always, then for a very long time. The very long duration of the epidemic in India and the large number of cases increase the likelihood of new mutations in the coronavirus, which can make existing vaccines ineffective and pose a threat to all mankind.

Knowing the optimal values of parameters, the corresponding SIR curves can be easily calculated with the use of exact solution [70, 72, 81] and compared with the pandemic observations before and after *T*_*c2*_. The results are shown in Figure by blue lines: *V(t)=I(t)+R(T)* – solid; dashed one represents the number of infectious persons multiplied by 5, i.e. *I* (*t*) ×5 ; dotted line shows the derivative (*dV* / *dt*)×100 calculated with the use of formula:

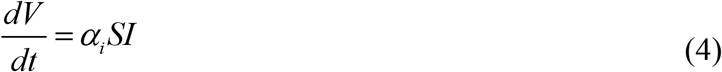

Equation (4) follows from (2) and (3) and yields an estimation of the real daily number of new cases. Red “Circles” and “stars” correspond to the accumulated numbers of cases registered during the period of time taken for SIR simulations *T*_*i2*_ and beyond this time period, respectively (all taken from Table 1). It can be seen that these values are in good agreement with the theoretical blue solid line.

**Figure.**
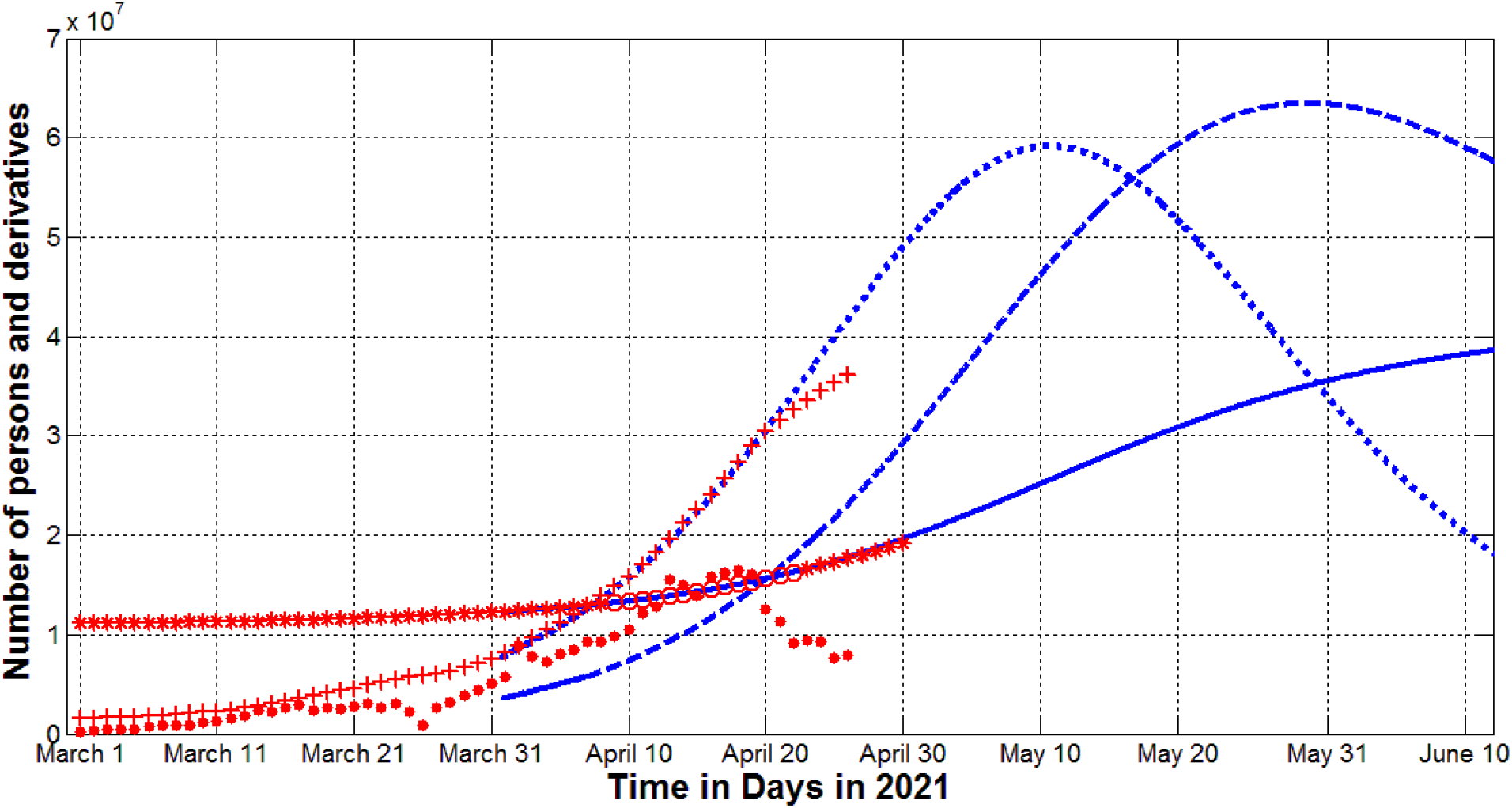
The COVID-19 epidemic dynamics in India. The results of SIR simulations are shown by blue lines. Numbers of victims *V(t)=I(t)+R(t)* – solid; numbers of infected and spreading *I(t)* multiplied by 5 – dashed; derivatives *dV/dt* (eq. (4)) multiplied by 100 – dotted. Red markers show the real number of cases and its derivatives: “circles” correspond to the accumulated numbers of cases taken for calculations (during period of time *T*_*c2*_); “stars” – number of cases beyond *T*_*c2*_ (all the values from Table1); “crosses” – the first derivative (6) multiplied by 100; “dots”-the second derivative (7) multiplied by 1000.

The blue dotted line shows that the daily number of the new cases will start to diminish after May 10, 2021, but the number of people spreading the infection *I(t)* will have its maximum only in the end of May, 2021. We can also compare the theoretical curve (4) with the average daily number of new cases which can be calculated with the use of smoothing registered *V*_*j*_ values [70, 81, 82]:

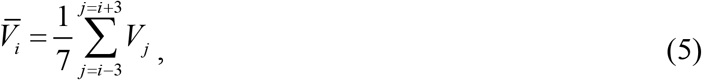

and its first derivative:

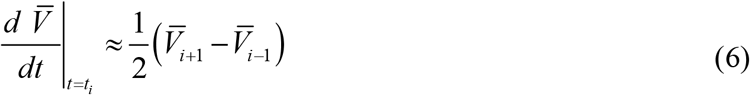

(see, e.g., [70, 81, 82]). The red “crosses” represent the results of calculations of the first derivatives (15) and are in a good agreement with the theoretical dotted line for moments of time before April 22, 2021.

## Discussion

Rather large differences between the real (15) and theoretical (13) values of the first derivative after April 22, 2021 can be explained by two factors. The first is related to changes in the dynamics of the epidemic. This is evidenced by the sharp changes in the second derivative of the average number of reported cases, which can be estimated using formulas:

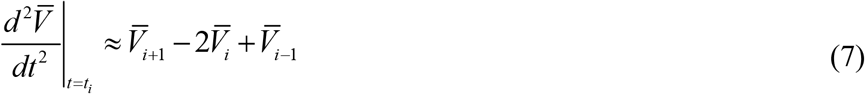

and (5) (see, e.g., [70, 81, 82]).

So we can talk about a new wave of pandemic in India, which may be weaker than the one we see in April 2021 due to the sharp decrease in the second derivative (7) after April 20, 2021 (see red dots in the figure). The course of the epidemic and new simulations will help to clarify these findings.

The second reason for discrepancies between formulae (4) and (6) may be the large number of unregistered cases observed in other countries [72, 78-80]. Estimates for Ukraine made in [72] showed that the real number of cases is about four times higher than registered and reflected in official statistics. Similar estimates can be made for the case of India. But the results already obtained show that the COVID-19 pandemic will be a problem for mankind for a very long time.

## Data Availability

Data is presented in the text

## Acknowledgements

The author is grateful to Dr. Vinod Varghese and Oleksii Rodionov for their help in collecting and processing data.

